# Acceptability of using mobile Health (mHealth) as an intervention tool for People with Drug Use Disorders in Tanga, Tanzania

**DOI:** 10.1101/2023.04.14.23288562

**Authors:** Castory Munishi, Harrieth P Ndumwa, Josephine E Massawe, Belinda J Njiro, Jackline Ngowi, Sanita Suhartono, Anja Busse, Giovanna Campello, Giovanna Garofalo, Pietro Cipolla, Cassian Nyandindi, Omary Ubuguyu, Bruno Sunguya

**Author notes:** **Email Addresses** CM HPN JEN BJN JE SS AB GC GG PC CN OU BS.

## Abstract

**Background:** With the increasing numbers of people with drug use disorders (PWDUD) in Tanzania as in other parts of the world the need for innovative interventions specifically tailored for this population has increased. Tanga, a coastal region on the Northeast of Tanzania has the second highest number of PWDUD in Tanzania. Evidence on the additional benefit in treatment and recovery process among PWDUD using digital health interventions is lacking. This study aimed to describe the acceptability of using a digital intervention to increase information access for PWDUD in Tanga region, Tanzania.

**Methods:** A cross-sectional descriptive study using both quantitative and qualitative approaches was conducted in Tanga Municipality and Muheza District. The quantitative approach used face to face interviews with a pre-tested questionnaire with 465 participants, while the quantitative method was carried out through In-Depth Interviews with 12 participants by the saturation point. Analysis was done descriptively to generate frequencies, cross tabulations, and chi-square test used to examine associations between categorical variables. Thematic analysis using codes was used to analyze qualitative data.

**Results:** Majority of the PWDUD 67.6% do not own mobile phones. Out of the 156 participants with mobile phones, only 6.4% owned a smartphone. Most of the participants, 83.6%, reported to live with someone who owns a mobile phone. Importantly, a significant number of participants, 98.5% from both areas showed readiness to use mobile phones to access information about the harmful use of substance and substance use disorder treatment options. Participants described how mobile phones can be useful to them in accessing information related to treatment and access to treatment options. The reasons they gave for not owning mobile phones included the need of money to buy drugs and the lack of money to buy credit drives them to sell their phones. A digital app called <u>Huru app</u> was developed during study as part of an information sharing campaign on substance use.

**Conclusion and recommendations:** The findings of this study helped to inform the target audience for the developed Huru app that should not be only PWDUD but the community at large. Despite the participants having expressed high readiness to use mobile phones to access drug use disorder treatment information, only few of them were found to own mobile phones but reported to live with family members who own mobile phones. Thus, a mobile phone intervention should also target their family members who are key in supportive treatment.

## Introduction

About a quarter of the 269 million people who used drugs at least once in their lifetime were from the African continent by 2020 (1). Both ageing and population growth are projected to increase the burden of mental health and substance use disorders by 130% in 2050 (2). Such unprecedented burden significantly increases the need for drug use preventive interventions. Tanzania harbor between 25,000 – 50,000 people who inject drugs (PWID) (3). Tanga, a Northeastern region of Tanzania is estimated to have around 25 hotspots of people with drug use disorder (PWDUD) with around 5000 males and 190 females (4). The number of PWDUD per 100,000 people aged 15 years and older in Tanga is reported to be 452, of which 47 are PWID (4). Due to unsafe injecting practices and engaging in unprotected sex, these people are at a higher risk of getting and transmitting Human Immunodeficiency Virus (HIV), blood borne infections such as Hepatitis C and other sexually Transmitted Infections (STIs) (5).

The increased use of mobile technologies such as smart phones has provided an opportunity for creating innovative solutions to address healthcare challenges (6). Digital interventions for drug use have been increasing at a quick pace over the past decade (7). They can complement practitioners-delivered interventions by offering information, monitoring and behavioral support to the clients. Further, they are often cost-effective with the potential to improve uptake, efficiency, and clinical effectiveness of the interventions (8,9).

Sub Saharan African countries like Tanzania are experiencing a large explosion of internet use where in the last decade the number of mobile subscribers and connections has quadrupled (10). Despite this massive potential, there is little utilization of digital interventions in Tanzanian healthcare (11). Such interventions may also be useful in addressing the gap in care and treatment of PWDUD in Tanzania. This study, therefore, aimed to describe the acceptability of using mobile health (mHealth) intervention among people who use drugs in Tanga region, Tanzania. The findings of this study may be useful in scaling up such efforts in addressing the growing burden of drug use, strained health system with fewer health workers to attend the clients, and high health care cost in the context of epidemiological transition in Tanzania.

## Materials and Methods

### Study Design and setting

The study employed a convergent mixed methods design using quantitative and qualitative approaches to determine the acceptability of using mobile health (mHealth) intervention among PWDUD in Tanga region, Tanzania from October 2020 to January 2021. A quantitative design was employed to obtain measurements such as the prevalence of mobile phone use with respect to demographic characteristics, whereas a qualitative design was employed to obtain a detailed understanding of the acceptability and feasibility of using mHealth among PWDUD. The study was conducted in two districts of Tanga Region, Muheza and Tanga City which were chosen to represent rural and urban settings respectively. Tanga is in North-Eastern part of Tanzania, bordering Kenya on the North and Indian Ocean on the East, it has a total population of 2,405,205 (12). It is reported to be the second city with the highest number of PWDUD in Tanzania (4). Muheza district has a population of 204,461 people (12). Tanga City is on the urban setting, located on the shores of the Indian Ocean in northeast Tanzania with a population of 273,332 as per 2012 census (12).

### Study Population and selection

The study included PWDUD with 18 years of age and above. PWDUD who were intoxicated or suspected to be under influence of drugs at the time of data collection were excluded. The main services available for PWDUD in Tanga are Methadone Assisted Treatment (MAT) at Bombo Regional Referral Hospital and privately-operated rehabilitation clinics and sober houses. In this study, participants were recruited from multiple sites, the streets harboring hotspots of PWDUD, rehabilitation clinics, sober houses and those who are on MAT.

### Sampling and sample size

Sample size was computed using Kish and Leslie proportion formula (13). A standard error of 5% and a proportion of 50% was used in estimating the minimum sample size as no similar studies have been previously conducted in Tanzania to assess the potential of deploying mHealth intervention among PWDUD. In this study, the minimum sample size computed was 465. Snowball sampling techniques were used to recruit study participants from different hotspots where PWUD are found in Tanga City and Muheza district. Members of civil society organizations involved in drug use rehabilitation and well oriented with the locations of PWUD in the two districts aided in reaching the study participants. Participants who met the inclusion criteria and consented to participate in the study were enrolled. For the qualitative study, we purposively chose six participants from each district to give further context on the responses obtained in the quantitative study and hence a total of 12 participants.

### Data measurements and tools

The structured interviewing forms were used to obtain data on the acceptability of using mHealth intervention among PWDUD in Tanga. The interviewing forms consisted of two sections, section one which was on the socio-demographic characteristics of the study participants and section two which was focused on the mobile phone usage of the study participants. It had a total of 22 questions, 20 close-ended and two (2) open-ended questions. The tool was developed in English language and then translated into Swahili; a national language spoken by majority in the country.

The socio-demographic characteristics included sex, age, level of education, literacy skills, and homelessness. The outcome variable was mobile phone possession which had sub elements which included the kind of mobile phone owned, whether they live with someone possessing a mobile phone, can they keep the mobile phone and for how long, tendency of selling mobile phones owned to get money for buying drugs and other needs, ability to purchase credit and whether they are willing to use mobile phones to access drug use disorder treatment related information and services.

A pilot study was conducted to pretest the tools before the actual data collection, using simulated participants who acted to have similar characteristics to the study participants. Necessary corrections and modification of the instruments were then made to obtain the final version that was used during the actual data collection.

### Data Collection

The study utilized both quantitative and qualitative approaches. The quantitative data was collected by the use of questionnaire, while qualitative data was collected through In-depth interviews. Information rich participants from identified during quantitative interviews were recruited for qualitative interviews in order to triangulate data obtained through quantitative methods and enable the researchers to get an insider perspective and context of mobile phone use among people with substance use disorder.

Quantitative data was collected by a team of 4 trained research assistants in each of the study districts. The collection took place from October to November 2020 simultaneously in both districts. The research assistants followed ethical principles in the conduct of the study and ensured that informed consent was given before starting to interview the participants. The interviews using questionnaires took about 30 minutes to complete. The interviews were conducted about 500 meters from the drug hotspot to ensure confidentiality of the respondent.

The qualitative approach through In-Depth Interviews explored the acceptability of using mobile mHealth intervention among PWDUD in Tanga to provide supplemental information on the quantitative findings. All interviews were conducted in Kiswahili and audio recorded with the permission of the study participants. Researchers applied the principle of bracketing to ensure that pre-understanding information do not influence the data. Field notes as a reflective diary was maintained for enhancement of reliability.

### Data Management and Analysis

Data collected through the interviewing forms was coded, entered in R statistical software version 4.0.0, and cleaned before the analysis was done. Numeric and categorical variables were analyzed using proportions for categorical variables, mean and standard deviation for numeric variables, and p-values to compare the findings in the two study sites. Socio-demographic information was summarized in tables and mobile phone use into both tables and graphs.

The qualitative data were analyzed through the thematic analysis framework to explore perceptions and acceptability of using mobile phones to access drug related information. The analysis involved six main phases of thematic analysis which included: familiarization with data, generating initial codes, searching for themes, reviewing themes, defining, and naming themes, and producing the report (14,15). Microsoft Word program was used to support data organization, coding, and themes identifications. In producing a report in this study, the data were presented as summaries and narratives and were illustrated with examples and quotations, capturing respondents’ perspectives and experiences.

### Ethical Consideration

Ethical clearance was obtained from the MUHAS Institutional Review Board (IRB), MUHAS-REC-12-2020-438. Permission to conduct the study was obtained from the Permanent Secretary of the Ministry of Health, the Regional Medical Officer of Tanga Region and the District Medical Officers from Tanga City and Muheza District. Permission to collect data from Bombo MAT clinic was further obtained from the Medical Officer in Charge and head of the MAT unit. Consent to participate in the study was sought from the eligible study candidates. No participant was allowed to participate without going through the consenting process.

## Results

### Socio-demographic Characteristics of the Study participants

A total of 481 participants were enrolled from both Tanga City and Muheza district, with the response rate of 100%. Three in four study participants were from the urban area (**Table 1**). The mean age was 37.7 ± 7.9 and was ranging between 20 and 72 years. Most participants were males 97.5% and single 54.1%. About 71% of the participants had attained primary level of education. More than three quarters of the participants 79.8%had a home with the majority 40.5% residing with their families. More than half of the participants 59.8% had used drugs for the first time at the age of 15 to 24 years, with a mean age (SD) of 24.3 ± 6 and, the majority 58.9% had used drugs for the period of more than 10 years (**Table 1**).

**Table 1:** Socio-demographic characteristics of the study participants N (%)

| Characteristic | Total<br>(N=481) | Muheza (N=121) | Tanga (N=360) | P-Value |
| --- | --- | --- | --- | --- |
| **Sex** |  |  |  |  |
| Male | 469 (97.5) | 121 (100) | 348 (96.7) | 0.09 |
| Female | 12 (2.5) | 0 | 12 (3.3) |  |
| **Age groups** |  |  |  |  |
| **Mean age (SD)** | 37.7 ± 7.8 |  |  |  |
| < 25 years | 24 (5.0) | 9 (7.4) | 15 (4.2) | 0.556 |
| 25-34 years | 182 (37.9) | 45 (37.2) | 137 (38.1) |  |
| 35-44 years | 202 (42.1) | 50 (41.3) | 153 (42.5) |  |
| 41+ years | 72 (15.0) | 17 (14.0) | 55 (15.3) |  |
| **Marital status** |  |  |  |  |
| Single | 260 (54.1) | 33 (27.3) | 83 (23.1) | 0.379 |
| Married | 91 (18.9) | 18 (14.9) | 73 (20.3) |  |
| Widowed | 14 (2.9) | 68 (56.2) | 192 (53.3) |  |
| Divorced | 116 (24.1) | 2 (1.7) | 12 (3.3) |  |
| **Education** |  |  |  |  |
| No Education | 23 (4.8) | 1 (0.8) | 22 (6.1) | 0.032* |
| Primary Level | 342 (71.1) | 85 (70.2) | 257 (71.4) |  |
| Secondary and Above | 116 (24.1) | 35 (28.9) | 81 (22.5) |  |
| **Can Read and Write** |  |  |  | 0.393 |
| Yes | 438 (91.1) | 113 (93.4) | 325 (90.3) |  |
| No | 43 (8.9) | 8 (6.4) | 35 (9.7) |  |
| **Has a Home** |  |  |  | 0.417 |
| Yes | 384 (79.8) | 93 (76.9) | 291 (80.8) |  |
| No | 97 (20.2) | 28 (23.1) | 69 (19.2) |  |
| **Residential Status** |  |  |  | 0.086 |
| Alone | 54 (12.9) | 15 (15.5) | 39 (12.1) |  |
| With Children | 16 (3.8) | 4 (4.1) | 12 (3.7) |  |
| With Family | 170 (40.5) | 27 (27.8) | 143 (44.3) |  |
| With Friends | 30 (7.1) | 10 (10.3) | 20 (6.2) |  |
| With Wife | 27 (6.4) | 9 (9.3) | 18 (5.6) |  |
| With Parents | 123 (29.3) | 32 (33.0) | 91 (28.2) |  |
| **Age First Used Drugs** |  |  |  | 0.211 |
| < 15 years | 19 (4.0) | 3 (2.5) | 16 (4.5) |  |
| 15 – 24 years | 287 (59.8) | 65 (53.7) | 222 (61.8) |  |
| 25 – 34 years | 156 (32.5) | 48 (39.7) | 108 (30.1) |  |
| 35 + years | 18 (3.8) | 5 (4.1) | 13 (3.6) |  |
| **Used Drugs for > 10 Years** |  |  |  | 0.150 |
| Yes | 282 (58.9) | 64 (52.9) | 218 (60.9) |  |
| No | 199 (41.1) | 57 (47.1) | 142 (39.1) |  |

**Table 2:**
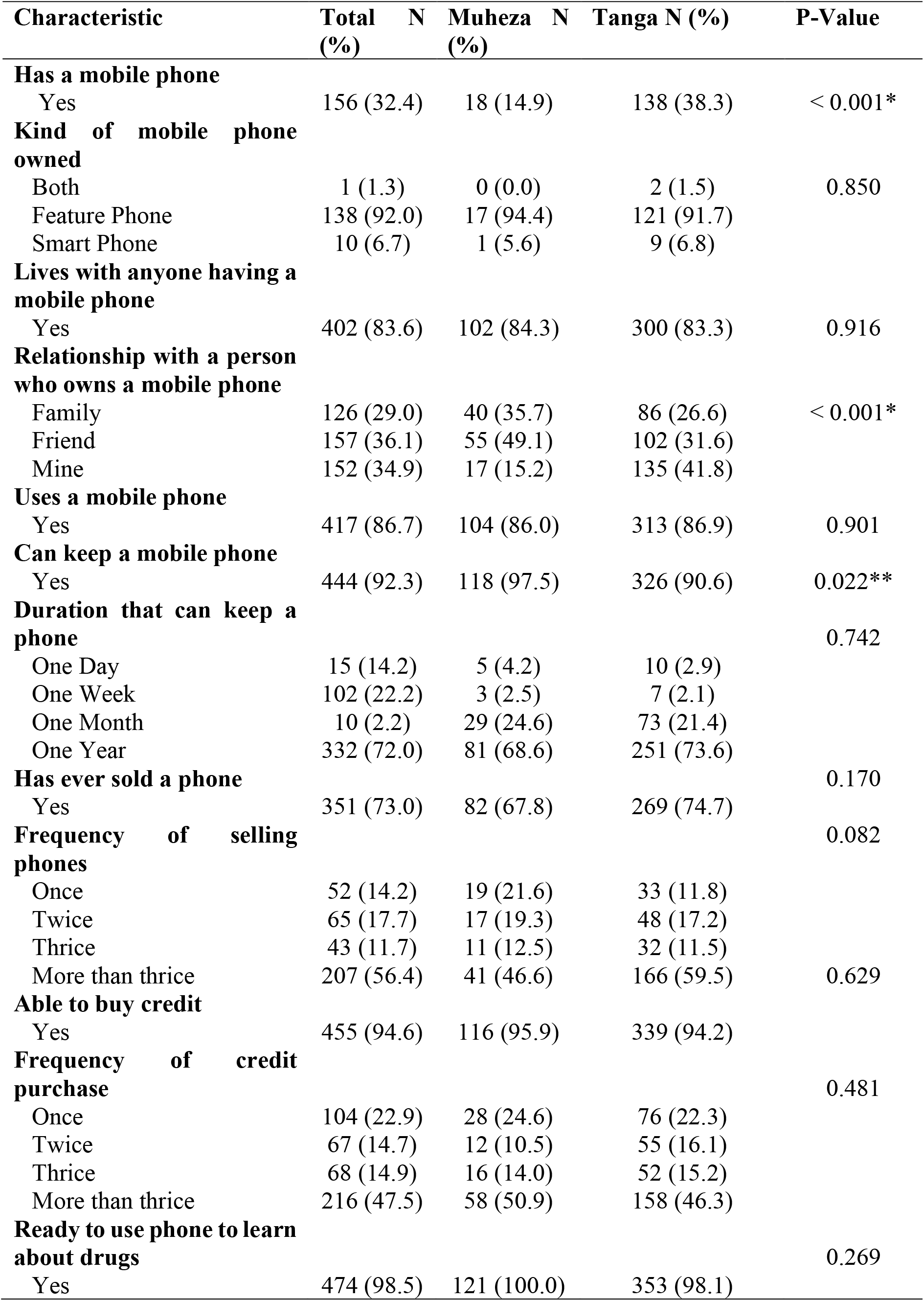
Mobile Phone Possession and Usage Patterns of the Study Participants

About a one third of the participants 32.4%reported to own mobile phones with more than two times higher prevalence of ownership in Tanga City as compared to Muheza (38.3% vs. 14.9%) (**Figure 1**). This shows a statistically significant association (p <0.001) between mobile phone ownership and urban residence.

**Fig 1:**
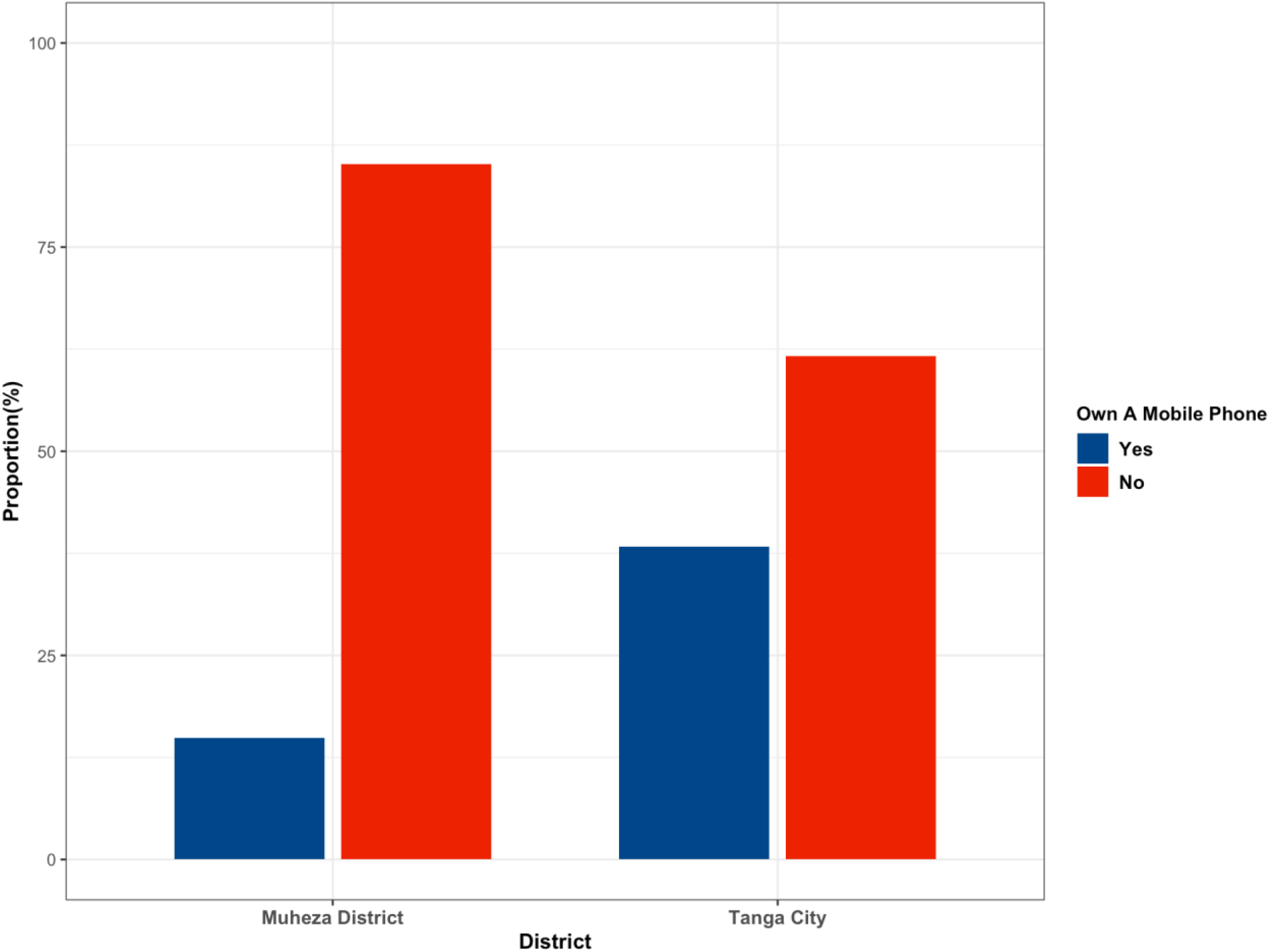
Comparison of Mobile Phone Ownership of Participants from Tanga and Muheza

Majority of the participants 92.0% owned feature phones compared to only 6.7% with smart phones ownership. Mobile phone ownership is further explored in qualitative design in which participants were interviewed about mobile phone ownership and the ability to maintain mobile phone ownership. Participants had mixed responses regarding mobile phone ownership, some stated that they do not own phones because it is expensive, as money is needed to buy credit. For example, the response from a participant in Muheza:

*“No, I don’t have because when you start using drugs it became so hard to get money for buying credit” (Participant No*.*08, Muheza)*.

Regarding the ability to maintain mobile phone ownership, quantitative findings revealed that about three quarters 73% have ever sold their phones when they lack money to purchase drugs and sustain their drug use. In the same regard over half 56.4% participants stated they have ever sold a phone they owned more than thrice. This is supported by qualitative findings in which a respondent stated that they sold their phone to get money for drugs:

“*I use phones but sometimes when I have problems, I sell it and use the money for drugs, I can buy another phone again when I get more money, but it has been a while since I used a mobile phone” (Participant No. 9, Muheza)*

In general, participants were aware of the importance of mobile phone use in communication, and they started to be using them for communication such as calling family and friends and chatting with their friends. Moreover, some participants were aware on the role of mobile phones in information sharing, like the use of the internet for accessing information. Examples of quotes from the participants:

*“Most of the time I use my phone for calling and chatting with my friends”* (*Participant No. 01, Tanga)*

*“Mobile phones are important in communication, to give us different information” (Participant No. 07, Muheza)*

*“I am watching YouTube videos on combating drug use” (Participant No. 03, Tanga)*

The majority of the participants, 83.6% had access to mobile phones and reported to be living with a family member, friend or a relative owning a mobile phone. It is also evident from the interviews as one of the participants said:

*“My mother, father and my uncle, they have mobile phones” (Participant No*.*07, Muheza)*

### The use of Mobile phones to access information of drug use

The majority of 94.6% of participants reported being able to purchase credit. Additionally, a significant number of participants, 98.5% from both areas have claimed to be ready to use mobile phones to access information about overcoming drug use. In Muheza district, all participants expressed readiness to use mobile phone to access information and for Tanga City 98.1% of the participants expressed readiness (**Figure 2**). This can also be evident from the interviews as participants were asked to respond on how they use their mobile phones and some of the responses were;

*“Mobile phones can give a lot of what you want; internet, you can learn about methadone, consequences of drug abuse and about relapse” (Participant No. 03, Tanga)*

*“…because we can get educated through mobile phones” (Participant No. 02, Tanga)*

**Fig 2:**
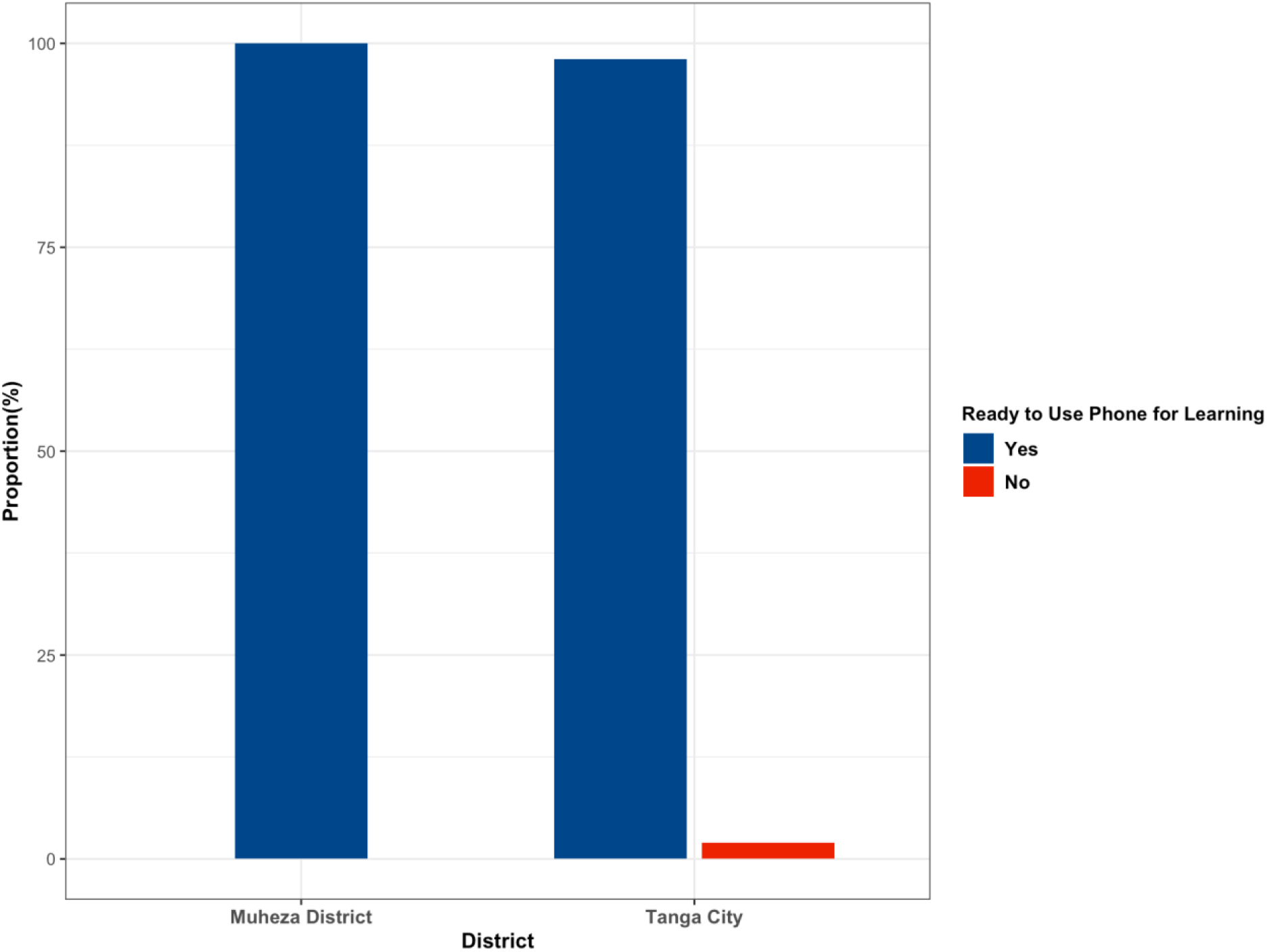
Comparison of Readiness to Use Mobile Phone for Learning about Substances of Participants from Tanga City and Muheza

Some participants indicated mobile phones can be useful for those who can read and write in accessing information but not for all, they suggested that there should be information centers in the streets which can offer free education to those who cannot read and write.

*“Yes, they can help especially for those who can read and write and those who can use mobile phones but for those who cannot be given information directly, also there should be centers or institutions where they will be provided with education” (Participant No. 11, Muheza)*

Another participant agreed that mobile phones could be useful, but went on further to state that they feel that there is the need of a human component in supporting their recovery. Even if there is a mobile app, there should be people who they can communicate well with and get to express their feelings and challenges. This shows that if there is a digital intervention for PWDUD it should be supported with a physical information center where people can call in through the app and they can also go directly to these centers to seek for further help.

*“Yes, it might be helpful though whether they will throw on the internet questions and write to us things that are like a lesson for us users, it would be better to communicate with us directly, because when someone stops drug use they need to belong somewhere as starting point of getting back to the community…*..*” (Participant No*.*04, Tanga)*

Some of the participant responses also indicated the importance of having small clips in the apps, they found that information through videos can be easy to grasp and understand but also they do not require one to be able to read and write in order to get the message.

*“Of course, when you have the apps and small clips, it can help as videos are interesting and they do not require you to be able to read and write to understand them” (Participant No. 04, Tanga)*

On the contrary, some participants expressed concerns regarding usefulness of mobile phones in helping them to quit drug use they felt that information through mobile phones may not be enough to help them quit they need more support like the human component which has been reported earlier.

*“That will be hard for me because you may give me information through the phone but sometimes, I may be in need of things that may help me stop, we need people to talk to and to support us to quit” (Participant No. 05, Tanga)*

Further some participants believed that drug use disorder treatment cannot only be provided through a mobile intervention, as treatment needs to be tailored to the individual’s needs. They thus think that in addition to the app, qualified health professionals, family members, peers and their communities are part of the whole support system to support the treatment and recovery of people with drug use disorders.

*“A mobile phone cannot make me stop using drugs because this is something that a person should decide from his/ her own heart that now I want to stop using drugs, but this is always hard due to the withdraw pains that a person get hence we fail to stop” (Participant No*.*09, Muheza)*

#### Huru App

*After successful completion of the study, a web and android application called [Huru app](https://huruapp.org/) was developed. The app was available to mobile phone and web users in Tanga City and Muheza districts as a complementary to the tradition Information, Education and Communication (IEC) materials during a campaign which was titled “Awareness in a caring community”*.

## Discussion

Two thirds of People with Drug Use Disorders in Tanga region did not own mobile phones, and only 10 participants were found to own smartphones. Almost three quarters of the PWDUD were found to be able to keep their mobile phones for more than a year. Importantly, PWDUD from both areas are also found to show readiness in using mobile phones to access information about the harmful use of substances and substance use disorder treatment options. A higher proportion of the participants (83.6%) reported to live with someone who own a mobile phone which helped to inform the intervention that was the outcome of this study that the intervention was designed such that the developed app can be used by PWUD along with community members to support them in accessing information about drug use disorder treatment and its facilities in Tanga and Muheza. Moreover, it was found that the app needs to run by people who are accessible to PWDUD and there should be information centers in the community where PWDUD can go and be supported in the recovery process.

The low proportion of mobile phone ownership among PWDUD in this study can be explained by the cost of mobile phone itself (16) but principally due to the consequences of drug use in most aspects of life, ranging from lack of education, unemployment, poor health, poverty, homelessness, and poor quality of life, making them not able to afford and maintain mobile phone ownership (17). These findings are contrary to those obtained in a study conducted in UK to access mobile phone ownership, usage and readiness to use by patients in drug treatment which found the majority (83%) of patients to be owning mobile phones (18). The prevalence is also significantly lower as compared to another study conducted in the US which found it comparable to ownership among US adults (19).

Additionally, the study found only about 10 participants to be owning smartphones contrary to another study which found a higher prevalence (57%) of smartphone ownership among people with substance use disorder (18). Higher prevalence of mobile phone ownership is observed among participants from the urban area as compared to the rural area (p < 0.001). This finding is also observed in other studies and is evidenced by the better social-economic activities, access to opportunities and the general economic status of the urban areas as compared to the rural areas (16,20). Moreover, the majority of the participants are found to be living with a family member, friend or a relative owning a mobile phone. This provides an opportunity to many PWDUD who do not own personal mobile phones to access digital health services and information on drug use, as they can be supported by family members owning mobile phones (21).

In this study about three quarters of the participants were found to be able to keep mobile phones for more than one year, however, the majority are observed to sell them frequently as evidenced from the interviews. Almost all the participants from both areas are also found to show readiness in using mobile phones to access information about substance use effects and treatment options. The use of mobile apps as an intervention for substance is still relatively a new concept, with the first app in the field of substance use targeted alcohol use and was developed around 2011 (22). A randomized trial to assess a smartphone application to support recovery from alcoholism showed that multi-featured smartphone application may have significant benefits to patients in continuing care for alcohol use disorder which is in line with what we have found in this study the app needs to go beyond information sharing (23). Given the novelty of this field, there is still more work to be done so as to develop working applications as the systematic review by Staigner et al. revealed that less than one third of the apps they studied showed mobile app users were in significantly better than the comparison conditions post treatment (24).

From this study and other studies as discussed, it is evident that, there is a potential of deploying mHealth interventions among people with drug use disorder in these respective study areas and even beyond. However, there are three things to consider in the deployment of the app. First, the app should be able to work in both smart phones and non-smart phones, second, the app should not only target PWDUD but the community at large which will be useful in helping PWDUD with no phones to access information, understand the impact of substance use and hence prevent other community members to start substance use and thirdly, there should be a supporting institution behind the app with accessible information centers in the community. Several studies have recommended the use of internet and computer-based forms of interventions to PWDUD as they have proved to have similar outcomes as compared to the traditional in-person approaches (25–27).

## Strengths and Limitations

A strength in this study is that it is the first study done in Tanga Region and Tanzania examining the potential of using mHealth interventions to increase information availability and connecting people with substance use disorders to appropriate services. The study employed a mixed method design which ensured gathering of sufficient evidence through triangulation. This study therefore lies groundwork for similar studies and provides information on how to design working digital health interventions for people with drug use disorders and their families in Tanzania. Further, through this study a digital health tool called HuruApp was developed.

Besides the strengths of this study, several limitations are noted. Due to stigma, the provision of socially desirable answers because of fear, may have resulted in response bias as substance use is a sensitive subject and is perceived illegal in Tanzanian communities. The study was only limited to two districts of Tanga region which were within the scope of the planned campaign on addressing substance use. Given limited time constraints, the qualitative component was not explored in full depths, it was added as an element to generate quantitative findings with context.

## Conclusion

A high proportion of PWDUD in Tanga own a mobile phone, an opportunity for integrating both physical and digital interventions in addressing substance use. This platform can be used to bridge knowledge and practice gaps towards substance use. In addition, the readiness to use such platform is high, and PWDUD showed readiness to use mobile phones to learn about substance use and substance use disorders. There is also an opportunity for the whole-community approach if targeted by information and messages aligning drug use knowledge, intervention, and harm reduction practices to address the burden of drug use in Tanga and areas with similar contexts. Evidence from this study enabled tailored mobile application development, the Huru app intervention has been developed to enhance sharing information and locating services for people with substance use. Studies to assess the effectiveness of this and other mHealth interventions in supporting recovery and treatment for PWDUD are warranted.

## Data Availability

Data used in this work is available in github for use.

https://github.com/castobary/huru_data_analysis.git

## Disclaimer

Anja Busse, Sanita Suhartono and Giovanna Campello are staff members of the United Nations. The views reflected in the article are those of the authors and do not necessarily reflect the views of the United Nations.

## Acknowledgement

We appreciate the support of the Muhimbili University of Health and Allied Sciences (MUHAS) staff for mentorship and logistics support. We further extend our gratitude to the Regional Medical Officer (RMO) of Tanga Region and District Medical Officers (DMOs) of Tanga city and Muheza district for their uncredible support during data collection. We appreciate the contribution of Dr Wallace Karata, Ms Irene Kaoneka, Mr Lupiana Hare, Mr. Said Bandawe and his team from the Gift of Hope Foundation for their support during field work in Tanga and Muheza. We would like to specially thank our research assistants and all study participants.

